# Index of potential contamination of urogenital schistosomiasis based on mass drug administration of praziquantel among school children in Benue State, Nigeria

**DOI:** 10.64898/2026.01.27.26345003

**Authors:** James Agada Okete, Faith Odije Okita, Eme Effiong Etta, Joseph Ele Asor, Benedict Onu Onoja

**Author notes:** Corresponding author’s.

## Abstract

Mass drug administration (MDA) of praziquantel is an intervention used in the treatment and prevention of schistosomiasis. Its effectiveness and sustainability require identifying subpopulations that are at risk of infection. A longitudinal survey was conducted among 3,810 subjects aged 5-19 years old recruited at baseline across ten council wards in Katsina Ala, Benue, Nigeria, to determine the prevalence, intensity, and index of potential contamination of urogenital schistosomiasis for three successive phases: three months, six months, and nine months post-treatment periods. Urine samples were processed using microscopy and reagent strips (Medi Test Combi 9). Prevalence of infection was recorded in all the phases of the surveys, with the first having the highest prevalence (12.30%), followed by the third phase (9.12%) and the second phase (7.60%), the difference being significant (P < 0.05). The highest intensity of infection (16 ova/10 ml urine) was observed in the first phase, followed by the third phase (15.10 ova/10 ml urine) and the second phase (11 ova/10 ml urine). Peak prevalence, intensity, and relative index of potential contamination (Rel. IPC) occurred among pupils between the ages of 10 and 14 years old in both sexes. The result of the relative contribution of each age group in polluting the snail habitat with *Schistosoma* eggs, thus enabling transmission, showed to a greater extent that children aged 10-14 years old were responsible for contaminating the environment with a bulk of *S. haematobium* eggs and for the transmission and maintenance of the disease in the area.

## Introduction

Schistosomiasis is a chronic and debilitating neglected tropical disease affecting poor rural communities in sub-Saharan Africa, where local populations seek water for cooking, drinking, washing, and bathing at schistosomiasis transmission sites (Abubakar *et al*, 2022). Over 280, 000 deaths per year in Africa is due to schistosomiasis, of which 150,000 and 130,000 deaths are due to urinary schistosomiasis and intestinal schistosomiasis, respectively. *Schistosoma haematobium* is a parasite that is responsible for urogenital schistosomiasis in different localities of Nigeria, particularly in the riverine areas where human-water contact activities are inevitable (Okete *et al*., 2021).

To encourage effective management of helminth diseases, including schistosomiasis, the 54th World Health Assembly (WHA) endorsed resolution 54.19 in 2001, which was to encourage member states to treat all school-aged children with praziquantel. Consequently, many sponsors, including NGOs and governments in endemic regions, are now paying keen interest to schistosomiasis and other neglected tropical disease (NTD) control (King, 2012). For instance, mass drug administration of praziquantel was implemented in Uganda and Tanzania in 2003 (Guyatt, 2010) and Nigeria in 2017 (Guyatt, 1999). According to Bowie *et al*. (2016), a cost-effective approach in the delivery of praziquantel through the schools was adopted in all these countries. However, elimination or reduction of the disease through this school-based method of treatment has been challenged by re-infection since other members of the community who may harbor the infection and potential sources of contamination or transmission are left out (Lemlem *et al*., *2010)*.

Nigeria is a signatory to the 54th World Health Assembly Resolution 54.19, which encourages affiliated states to treat 75-100% of all school-aged children, pregnant women, and other vulnerable groups by 2020 (Schistosomiasis Control Initiative, 2020). Interestingly, part of the mandate of this resolution, which was praziquantel mass administration in schools, has commenced fully in many states of the federation. To strengthen this objective, estimation of prevalence and intensity and the index of potential contamination of urogenital schistosomiasis in areas where mass drug administration of praziquantel has been implemented is key. This would ascertain the age groups that are contributing the bulk of Schistosoma eggs, thereby polluting the environment so that treatment can be implemented in a cost-effective manner.

This survey was conducted in school-aged children because they are a highly vulnerable group, given the importance of the microbiome to them (Célia *et al*., 2024). Many studies have assessed what factors impact the composition, abundance, and diversity of the prevalence and intensity of schistosomiasis in school children but have yet to evaluate the IPC, and thus, they should be the first targeted group for intervention because of the detrimental effects the disease has on their growth and development (Afiukwa *et al*., 2019; Jozef *et al*., 2001). Furthermore, they represent the infection status in the population, as they consistently have the highest prevalence and transmission of schistosomiasis, and treatment via schools is also feasible and cost-effective (Bowie *et al*., 2004).

Index of potential contamination (IPC) is the most powerful predictive methodology and relevant epidemiological measure to identify the relative potential of different age groups at higher risk of infection and responsible for the transmission and spread of this disease in a particular area than the use of prevalence and/or intensity (Jozef *et al*., 2001). The identification of risk factors for *S. haematobium* infection contributes to a better understanding of the transmission process and the definition of control programs in particular localities (King *et al*., 2004).

There is increasing attention given to the dilution effect of biodiversity on infectious diseases. For example, the presence of predators reduces anti-cercarial behavior in tadpoles, contributing to the increase in exposure rates of tadpoles to trematode parasites (Shun Zhou et al., 2024).

Although several epidemiological studies for intestinal schistosomiasis due to *S. haematobium* infection were carried out in different parts of Nigeria, none of them yet estimated the IPC of this disease (Eigege, 2017 and Eigege, 2020). Moreover, there is no updated information on the infection profiles and transmission potential of urogenital schistosomiasis of the current study site, except for a study conducted a few years ago by Okete *et al*. (2019).

However, a health strategy for the attainment of effective parasitic disease control programs demands knowledge on the magnitude of the disease and its changes over time, with ecological, cultural, behavioral, and other factors. Therefore, the present study aimed to provide epidemiological data on the magnitude, intensity of infection, and potential transmission of *S. haematobium* infections among school children in different council wards in Kastina-Ala, Benue State, Nigeria.

## Materials and Methods

### Description of the study area

The study area was Katsina-Ala Local Government Area (Figure 3). Katsina-Ala is one of the largest Local Government Areas in Benue Sate. It has an area of 2,402km^2^ and a total population of 224,718 people (2006 Census). According to Omudu and Iyough (2017), the ethnic capital of Katsina-Ala Local Government Area is Katsina-Ala, which has the Tiv and Etulos as the indigenous people. However, there are other ethnic groups, including the Akweya and Nifon, Jukun, Hausa, Igbo, and Igala people, that are residing in the area.

Katsina-Ala Local Government Area lies within the lower River Benue trough in the middle-belt region of Nigeria. It lies between longitude 7.33 ^□^ E and 9.0 ^□^ E east of the Greenwich meridian and latitude 7.33 ^□^ N and 9.05 ^□^ N north of the equator and borders five other Local Government Areas of Benue State: Ukum (North), Logo, Kwande (South), and Ushongo (West) (Houmsuo *et al*., 2011). It experiences two weather conditions: “a warm, humid, rainy season and a blistering dry season. In between the two seasons, there is a brief harmattan, occasioned by the west-east trade winds, with the main feature of dust haze, intensified by coldness and dryness.” The rainy season begins from April to October with an annual rainfall in the range of 100-200 mm when the temperature reaches 28-30°C, and nighttime hovers around 24-25°C. In the dry season, the daytime temperature can drop to 19°C. According to Houmsuo *et al*. (2011).

Katsina-Ala is a cosmopolitan settlement on the north bank of the river from which the town takes its name. The people of Katsina-Ala Local Government Area are predominantly farmers. Over 75% of the population engages in agriculture, making agriculture the mainstay of the economy of the people.

**FIG 1.**
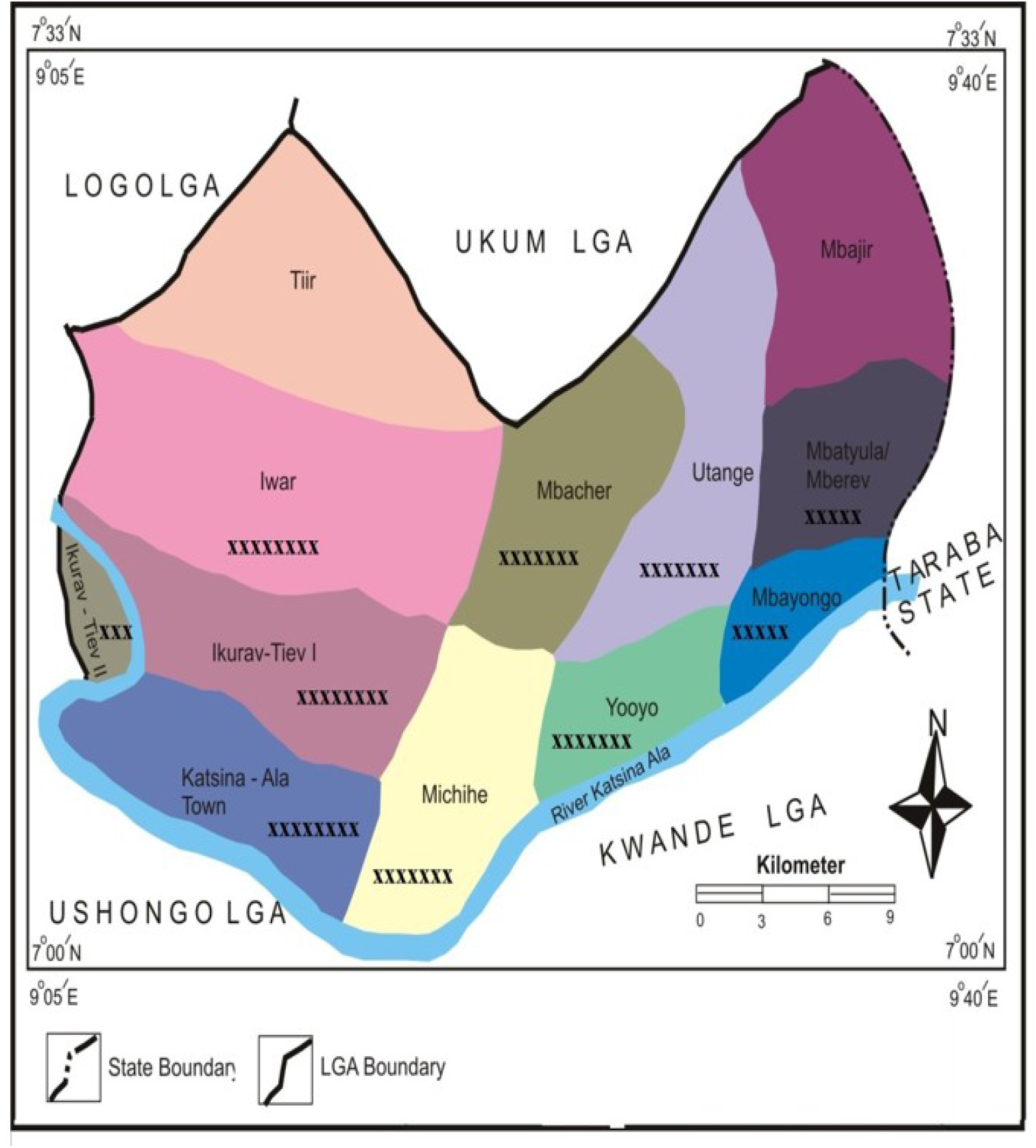
Map of Katsina-Ala showing the study areas (Modified from https://www.onlinenigeria.com) **Key:** xxxxx = Council wards where samples were collected.

### Study design and sample size determination

This study employed longitudinal survey methods, which were conducted in three successive phases with the same pupils (aged 5-19 years old) surveyed at each point in time (Omar and Bruno 2016). The sample size for this study was calculated using the normal distribution formula (N=□^2^ pq/i^2^) where

N = the desired sample size

□= the normal Z score corresponding to the risk of error α=5%

p = baseline prevalence of schistosomiasis in the study area (40.1%) (CDC, Benue state chapter preliminary report, 2018)

q = 1-p; I = the precision, fixed at 5%.

N = (1.96)^2^ (0.401) (1-0.401)/0.05^2^) = 363.

We considered the cluster effect, such that children from the same community attending the same school could acquire an equal risk of the disease and expected attritions. Hence, N (363) was multiplied by 5% and added to the sample size, N, to approximate the baseline sample size to 381 for each council ward. Therefore, the total sample size at baseline was 3,810 schoolchildren.

### Inclusion criteria for selection of participant communities and schools

Participant communities and schools were selected based on the baseline prevalence and praziquantel mass treatment reported by the CDDs/NTDs unit of the Federal Ministry of Health. Among other factors, the key factors that influenced the selection of participant communities and schools include the safety of the communities at the time of this research, epidemiological factors for transmission of schistosomiasis, such as proximity to fresh water bodies, and records of *Schistosoma* infections reported by other researchers. Before commencement of the main study, we first visited the Local Government Area to be familiarized with the people and to ascertain some key epidemiological factors (presence of freshwater bodies, Presence of freshwater snails, and proximity of schools to these freshwater bodies).

### Ethical approval and informed consent

Ethical Clearance (Ref. No: MOH/STA/204/VOL.1/71) (Appendix 2) used during this study was granted by the Research and Ethics Committee unit, Benue State Ministry of Health & Human Services (MOH).Verbal consent and permission to conduct the survey in primary schools were obtained from the Education Secretary, Katsina-Ala Local Government Areas, based on an introductory letter (Appendix 1) and ethical clearance. The introduction letter was issued by my supervisors on behalf of the Departmental Research Ethics Committee. The research was strictly conducted in line with ethical guidelines recommended by WHO (2011) with little modification as stipulated in the ethical clearance guidelines issued to us by the Benue State Ministry of Health & Human Welfare.

### Reconnaissance survey

Before the commencement of the research and we took a familiarization tour of all the schools and communities that were chosen to obtain their full consent. We met with the heads of school and community heads to address the reasons for doing the work and its potential benefits. The head teachers were then asked to inform the pupils and others in their respective villages/schools about the survey. In the schools, the volunteers were identified by raising their hands, after which they were given forms to fill. These consented pupils were registered and used as study subjects.

### Praziquantel administration

The Federal Ministry of Health provided the praziquantel through the Centre for Disease Control and Prevention (CDC), Benue State Chapter. Mass administration of the drug in a standard single dose of 40 mg/kg according to body weight to the respective schools/schoolchildren was done by the Local Government Area NTD Coordinator, assisted by some schoolteachers.

### Post-treatment survey

Three successive surveys were conducted following mass administration of praziquantel. The first survey was conducted in April 2018 and was repeated after every 3 months up to November 2018.

### Urine sample collection for parasitological assessment Measurement of haematuria and proteinuria

After responding to the questionnaire, a transparent sterile universal bottle was labeled according to the number on the questionnaire and given to each respondent to produce 15 ml of his/her own midstream urine. Urine specimens of consented subjects were collected after a 20-30 minute brief physical exercise between 10:00 am and 2:00 pm (Cheesbrough, 2005).

Each of the urine samples collected was first examined for appearance and then separated into two sub-samples. The appearance of urine was classified into four categories: clear, cloudy yellow, cloudy brown, and bloody red. Macro- or gross haematuria was diagnosed if the urine was cloudy brown or bloody red (Red Urine Study Group, 1995).

One part of the sub-samples was used immediately for assessment of micro-haematuria and proteinuria in the field using Medi-Test Combi 9 (Macherey-Nagel Eurl GumbH & Co. KG. Germany) reagent strips in line with the procedures of WHO (2010). The reagent strip was dipped into the urine, making sure all the spots were immersed. After approximately 3 minutes, the strip was removed from the urine container, drawing it across the rim of the container to remove excess urine. The strip was held horizontally to prevent the chemicals from the different test spots from being mixed together. After 30 seconds, the test strip was compared with the colour scale on the reagent strip container to record the protein and blood content (Appendix 4). The protein content was recorded as negative (-) or positive; + (30 Mg/dl, ++ (100 Mg/dl) or +++ (500 Mg/dl) while the content of the blood was recorded as negative or positive; + (ca. 10 Ery/µl), ++ (ca. 50 Ery/µl), or +++ (ca. 250 Ery/µl) respectively.

### Laboratory determination of Prevalence and intensity of urogenital schistosomiasis

The other sub-samples of urine were preserved with two drops of 40% formaldehyde (WHO, 1985; WHO, 2008; Bassey, 2012), tightly covered, labelled, and transported to Benue Diagnostic Laboratory, Katsina-Ala, for a quantitative parasitological examination. Laboratory examination of urine samples using sedimentation methods was carried out in line with the procedures outlined by Cheesbrough (2005) and employed by Adie *et al*. (2015), Ekpo and Mafiana (2008), Houmsou *et al*. (2011), and Lengeler *et al*. (2000). Ten (10 ml) of each urine sample was transferred into a 15-ml conical centrifuge tube and spun at 350 rpm for 15 minutes. The supernatant was decanted manually, leaving about 0.5 ml of the fluid with the sediment at the bottom of the tube. The sediment was thoroughly mixed, and then a drop of the mixture was transferred to a microscopic slide. A drop of 40 % Lugol iodine was added to stain the eggs if present, and the slide was covered with a coverslip and examined under a light microscope using x 10 objective lenses. The eggs of Schistosome *present* were identified by their distinctive characteristics, as seen under the microscope and recorded in confirmation of the result of reagent strips as positive for prevalence study, while the actual egg count was done per 10 ml of urine for intensity study. Based on the number of egg present per 10 ml of urine, urine samples were classified as follow: 0 egg per10 ml - no infection; 1-49 eggs/10 ml-light infection; 50-199 eggs /10 ml - moderate infection and 200 and above eggs/10 ml - heavy infection (WHO, 1995; Red Urine Study Group, 1995).

### Calculation of Prevalence, intensity and relative index of potential contamination re infectivity

Prevalence/intensity and relative index of potential contamination was calculated as described by Chitsulo and Engel, 2004) and employed by Tadesse and Tsehaye (2010) Prevalence of infection was calculated and expressed as a percentage with the formula below:

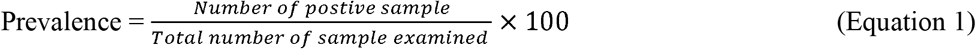

Intensity = Number of eggs per 10 ml of urine.

The relative index of potential contamination was calculated with the formula below: Rel.Indexofpotentialcontamination

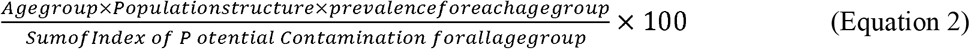

### Statistical analysis

Data were entered into Microsoft Excel 2007 and exported into a computer-based software programme, SPSS version 17.0 (Statistical Package for Social Sciences), and analyzed using appropriate statistical tools. The chi-square (χ^2^) test was used to determine the significance of the association between:

Schistosomiasis level varies between locations, age, and sex.

Analysis of Variance (ANOVA) was used to determine significant differences in the prevalence of infection among the different Council Wards based on the different phases of surveys.

## Results

### Prevalence of urinogenital schistosomiasis by post-treatment survey phases and Council Wards

Three months after mass administration of praziquantel, a total of 3,810 pupils from different council wards of Katsina-Ala were examined for urogenital schistosomiasis. Infection occurred in all the Council wards during the three phases of post-treatment surveys except in Michihe Council wards, where there was zero prevalence in the last phase (nine-month post-treatment survey) (Table 5). Eggs of *S. haematobium* were found in all the samples except in a few cases where there was mixed infection of S. *haematobium* with *S. mansoni* (Plate 1).

**PLATE 1 :**
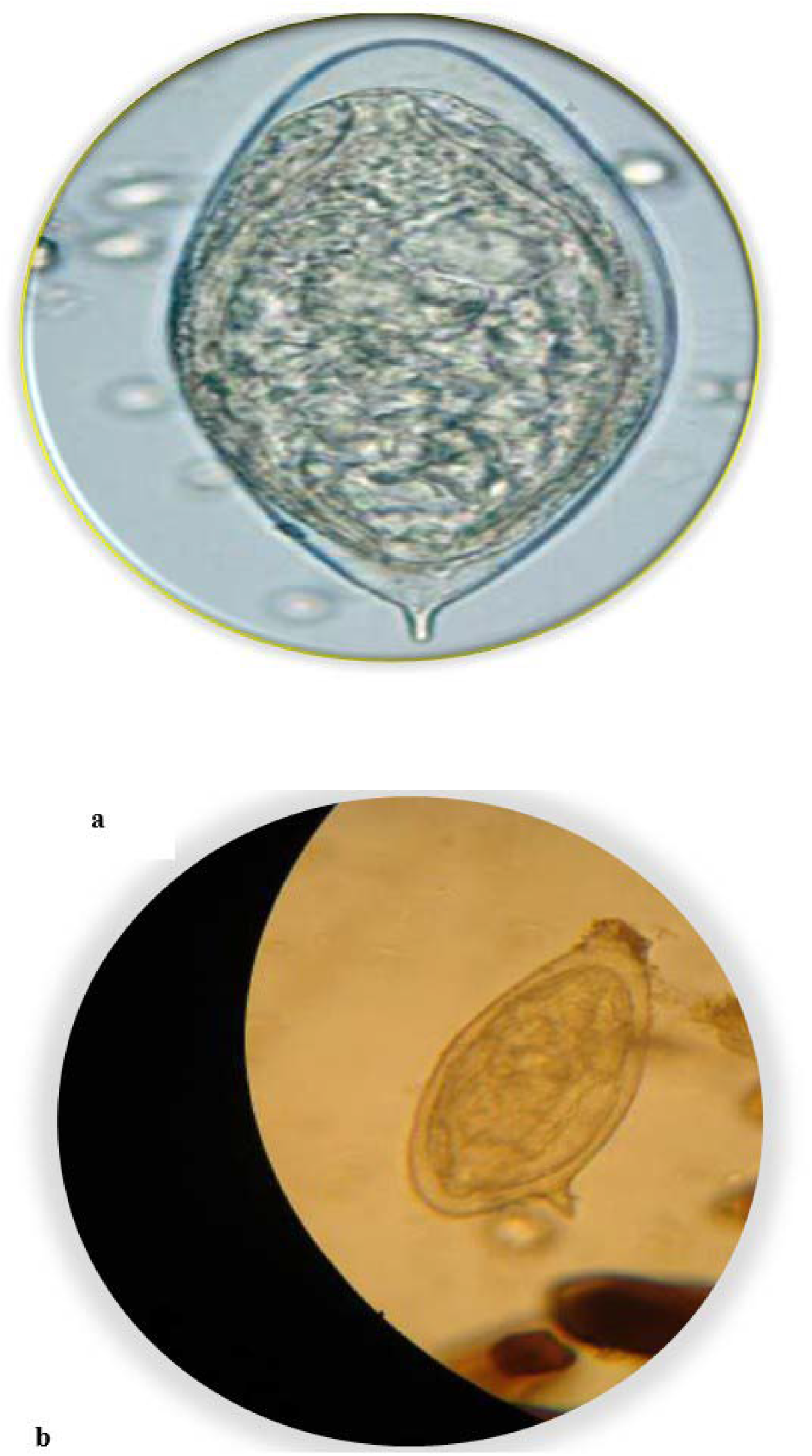
Eggs of schistosomo found in urine of pupils: (a) *S. haematobium;* (b) *S. mansoni*.

The overall prevalence was higher in the three-month and nine-month post-treatment surveys (12.30 % (471/3810) and 9.2% (335/3637) than in six month post treatment surveys, with an infection rate of 7.6 % (275/3,612). There was no significant difference (P > 0.05) in prevalence among the different phases of the surveys.

### Prevalence of urinogenital schistosomiasis by age and sex

The prevalence of urogenital schistosomiasis among pupils after a 3-month interval post-treatment survey is shown in Table 6. In the survey, 3,810 pupils (2,447 males and 1,363 females) aged 5-19 years old were examined. Of the pupils infected, 14.0% (343/2,447) were males, while 9.4% (128/1,363) were females. There was a significant difference (χ^2^ = 6.64, df = 1; P < 0.05) in prevalence between the sexes. For males, the age-related prevalence was as follows: 5-9 (9.52%), 10-14 (16.58%), and 15-19 (12.52%), while among females, prevalence of 6.78%, 8.93 % and 10.78 % was recorded for the age groups of 5-9, 10-14 and 15-19 years old respectively. The overall prevalence of infection was 12. 36 %.

In a 6-month post-treatment survey, a total number of 3,612 pupils (2,112 males and 1,500 females) were examined. 8.46% (66/780) of males and 5.42% (30/554) of females were infected, the difference being significant (χ^2^ = 6.64; df = 1; P < 0.05). Pupils of all ages were infected, the highest prevalence (10.01%) being recorded among pupils of the age group 10-14 years old in males, while among females, the highest prevalence (5.51 %) was recorded in the age group 5-9 years old. The overall prevalence of infection was 7.20 %.

In a 9-month post-treatment survey, 3,637 pupils (2,125 males and 1,512 females) were examined. 10.16% (216/2,125) of males and 7.87% (119/1,512) of females were infected. The difference in prevalence between the sexes is significant (χ^2^ = 6.64, df = 1; p < 0.05). For males, the age-related prevalence was as follows: 5-9 (9.16 %), 10-14 (11.21 %) and 15-19 (9.50 %) while among females, prevalence of 10.30 %, 7.75%, and 7.70 % were recorded for the age groups of 5-9, 10-14 and 15-19 years old respectively. The overall prevalence of infection was 9.21%.

### Relative index of potential contamination (Rel. IPC)

The relative contribution of each age group in polluting the snail habitat with *Schistosoma* eggs, thus, enabling transmission, is often expressed as the Relative Index of Potential Contamination (Rel. IPC) is shown in Figure 7. For the 3 months post-treatment survey, the age groups 5-9, 10-14, and 15-19 years old had the following Rel. IPC: 15.45%, 61.32%, and 23.22%, while in the 6^th^ month post-treatment survey, the age groups 5-9, 10-14, and 15-19 years old had the Rel. IPC values of 14.48%, 61. 85%, and 23.67%, respectively. In the 9^th^ month of post-treatment survey, the contribution of pupils in polluting snail habitat by different age groups was as follows: 5-9 (14.06%), 10-14 (63.38%), and 15-19 years old (22.55%).

**FIG 2.**
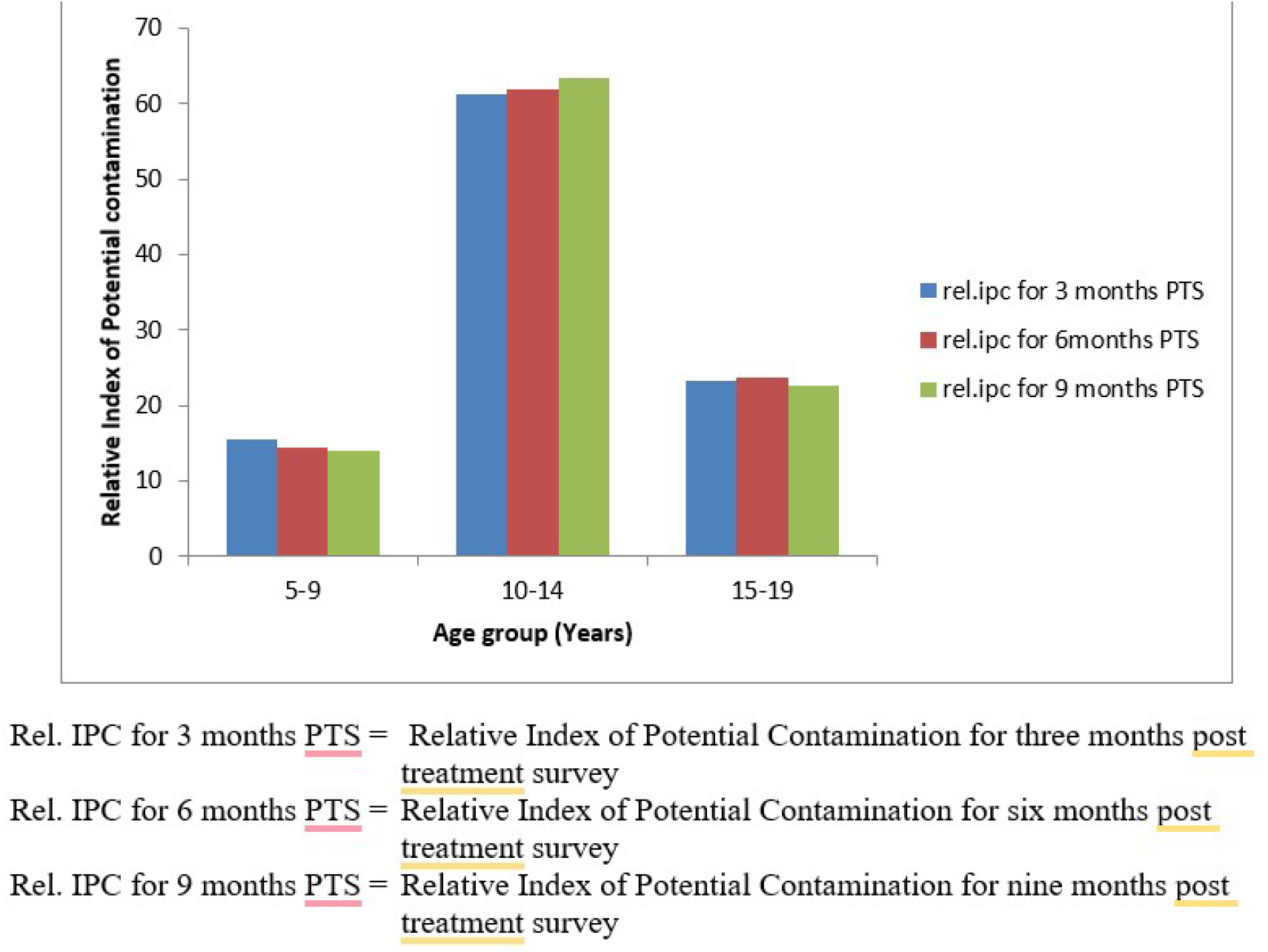
Relative index of potential contamination (rel.ipc) by age groups of pupils based on one round of mass administration of Praziquantel

## Discussion

Monitoring and evaluation of the performance of praziquantel mass drug administration and advocacy campaigns for schistosomiasis control, such as coverage and evaluation of impact on prevalence and intensity as well as the index of potential contamination, are necessary components of schistosomiasis control and intervention (Ben and Useh, 2017).

Investigations showed that Mbatyula Council Ward was the most endemic area for urogenital schistosomiasis despite one annual mass administration of Praziquantel with a prevalence of 25.05 %, 14.9 %, and 14.9 % in the 3 months, 6 months and 9 months’ post-treatment survey, respectively. Prevalence of urogenital schistosomiasis has been reported by several researchers in Nigeria: Bassey (2012) recorded a prevalence of 46.17 % among people in FCT; Adamu (2001) showed a prevalence of 47 % in Oju Local Government Area of Benue State, Nigeria, while Houmsou *et al*. (2011) recorded a prevalence of 56.30 % in Buruku Local Government of Benue State; Asor and Arene (2010) reported a prevalence of 70.1 % among inhabitants of Odau community in Rivers State;

The low prevalence recorded in this present study, when compared with the above-mentioned studies, may be because treatment was administered to the subjects before the commencement of the study. It was also observed that prevalence in the first (3 months post-treatment) survey was higher than that of the second (6 months post-treatment) survey, which, in turn, was lower than the third (9 months post-treatment) survey (Table 1). This may probably be due to re-infection after treatment, since contact with schistosome cercaria-infected water was not abated by the subjects. This agrees with the reports of Emily *et al*. (2022), who reported that praziquantel treatment only reduces parasitaemia, but does not prevent its reinfection.

**Table 1:** Prevalence of urinogenital schistosomiasis after a 3-month interval post-treat|nent survey in different council wards in Katsina-Ala. Benue State, Nigeria.

| Council<br>Wards | Post-treatment survey periods (months) |  |  |  |  |  |
| --- | --- | --- | --- | --- | --- | --- |
|  | Three months |  | Six Months |  | Nine Months |  |
|  | No.<br>Examined | No. infected | No.<br>Examined | No. <u>infected</u> | No.<br>Examined | No. <u>infected</u> |
| Iwar | 340 | 23 (6.8) | 325 | 15 (4.6) | 340 | 30 (8.8) |
| Ikurav-tiel I | 226 | 30 (13.3) | 220 | 20 (9.1) | 221 | 27 (12.2) |
| Mbatvula | 257 | 64 (25.0) | 215 | 32 (14.9) | 215 | 32 (14.9) |
| Mbacha | 220 | 45 (20.5) | 202 | 25 (12.4) | 202 | 25 (12.4) |
| Michile | 310 | 71 (23.0) | 301 | 30(10.0) | 310 | 0 (0.0) |
| Kastina-Ala | 1103 | 102 (9.2) | 1,065 | 80 (8.0) | 351 | 35 (10.0) |
| Yoooyo | 385 | 62 (16.10) | 351 | 27(8.0) | 351 | 35 (10.0) |
| Utange | 205 | 19 (9.3) | 200 | 18 (9.0) | 200 | 27 (13.5) |
| Ikurav-teil II | 330 | 23 (7.0) | 318 | 16 (5.0) | 318 | 35 (11.0) |
| **Total** | **3,810** | **471(12.3)** | **3,612** | **275 (7.6)** | **3,637** | **335 (9.20)** |

The study also showed that both prevalence and intensity were gender- and age-dependent. In relation to gender, males recorded a higher prevalence than females in all three post-treatment surveys (Table 2). This phenomenon is similar to the reports of Aribodor *et al*. (2019) in Enugu State, Abdullahi *et al. (*2011) in Kwara State, and Anosike *et al. (*2003) and Anyanwu *et al*. (2002) in Abia State, who recorded higher prevalence in males than females. Work by other researchers in other parts of the world also confirmed this pattern. This may be due to the fact that males showed more water contact than females. This agrees with the work of Etim *et al*. (2012), who reported a prevalence of 54% in males and 30.0% in females, and Asor and Arene (2010), who reported a prevalence of 73.3% for males and 66.7% for females in the Odau community of Rivers State, Nigeria. However, this was in contrast with that of Abdoulaye *et al*. (2015) and Amali (1988), who found prevalence in females to be higher than in males.

**Table 2:**
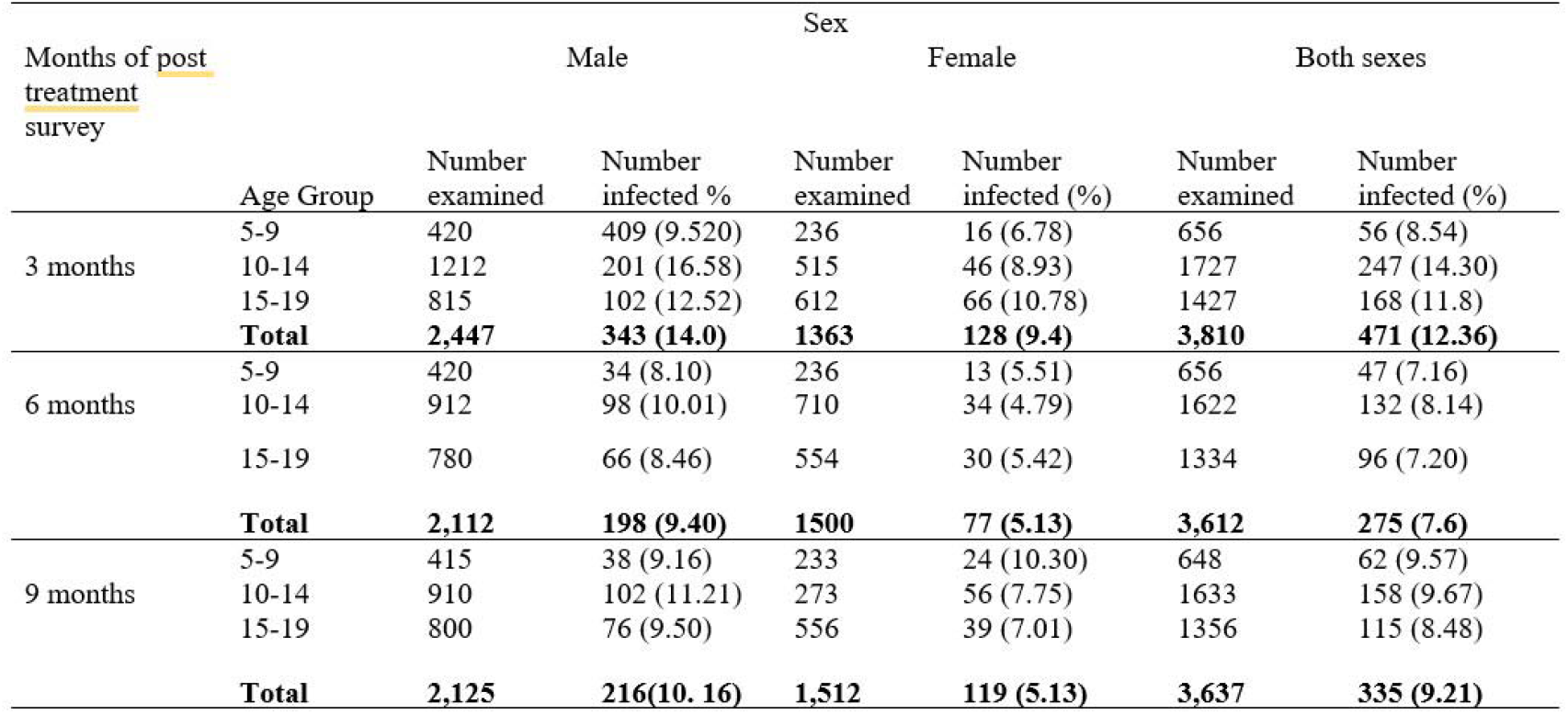
Prevalence of urinoginatal schistosomiasis after a 3-months interval post treatment survey based on sex and age of children.

The age group 10-14 had the highest prevalence and intensity of infection. This could be as a result of the group constituting a higher proportion of the population and also being responsible for the highest relative index of potential contamination of water. Bassey (2012), Asor and Arene (2010), and Asor (1995) also found similar observations, where the age group 10-14 years old constituted the most in polluting the environment, as they had the highest index of potential contamination. The high prevalence may also be due to the biophysiology of this group that favours the survival of the parasite in their body.

The relative index of potential contamination (Rel. IPC) is the relative contribution of each age group in polluting the snail habitats with *Schistosoma* eggs, thus enabling further transmission of the disease (Ekanem *et al*., 2017). Both Rel. IPC and re-infectivity are important components of control interventions that need careful evaluation if transmission must be interrupted. In this present study, it was observed that all the age groups were potential contributors to polluting the snails’ habitats. The highest potential contributor is being recorded among the age group of 10-14 years old. Although this observation is at variance with the findings of Jean and Keiser (2018), who reported that the age group 5-9 years old harbours the highest Rel. IPC. This difference may be as a result of the fact that their investigation was a community-based survey (CBS) and concentrated more on preschool and out-of-school-aged children, while our studies were school-based surveys (SBS).

The results obtained for Rel. IPC based on the groups is not far from that of the prevalence of the disease based on age groups, in which the highest prevalence was recorded in the age group of 10-14 years old. The reason for the observation has already been discussed under the prevalence of infection

## Conclusion

The present study has clearly indicated the prevalence of *S. haematobium* infection in the study area and reported two new infection foci. IPC values revealed that children aged 10-14 years were mostly impacted by heavy egg burden and take the higher position in contamination of the environment with the eggs of *S. haematobium*. They also represent the main transmission group and the most important age groups in maintaining the disease in the locality. Thus, the age group should be targeted for control and treatment. Water supply and sanitary conditions of the society should be maintained through education and awareness creation to reduce the overall worm burden. Further study incorporating snail surveys and awareness level of the community should be conducted to ensure effective and sustainable control of schistosomiasis in this area.

## Supporting information

Ethical Clearance

## Data Availability

All data produced in the present work are contained in the manuscript.

## AUTHORS’ CONTRIBUTIONS

**James Agada Okete: C**onceptualized, designed, and managed the data of the study.

**Faith Odije Okita:** Undertook the fieldwork and collection of data.

**Eme Effiong Etta:** Performed data analysis and interpretation.

**Joe Ele Asor:** Carried out data visualization, validation, and supervision.

**Benedict Onu Onoja:** Prepared the original manuscript, edited and submitted it (Corresponding Author).

## AUTHOR APPROVALS

All authors contributed to the development of the final manuscript and approved its submission. Also, this manuscript has not been accepted or published anywhere.

## APPENDIX I

**LIST OF GLOSSARIES**

1. **Advocacy Campaigns**: Organized efforts to raise awareness and influence policies or behaviors regarding a health issue, such as promoting treatment or prevention of schistosomiasis.
2. **Anti-Cercarial**: Refers to substances or measures used to kill or prevent infection by cercariae, the larval stage of schistosomes.
3. **Fresh Water**: Naturally occurring water with low concentrations of salt, found in rivers, lakes, and ponds—often associated with schistosomiasis transmission.
4. **Haematuria**: The presence of blood in the urine, a common symptom of urogenital schistosomiasis.
5. **IPC (Infection Prevention and Control)**: Policies and practices aimed at preventing the spread of infections, especially in healthcare or community settings.
6. **Mass Drug Administration (MDA)**: A public health strategy where an entire population or targeted group is given medication (e.g., praziquantel) to control or eliminate diseases, regardless of infection status.
7. **Medi-Test Combi 9**: A type of urine dipstick test that detects multiple parameters (e.g., blood, protein, glucose) to aid in diagnosing infections or kidney problems.
8. **Microbiome**: The collection of microorganisms (bacteria, viruses, and fungi) living in a particular environment, such as the human body or a water source.
9. **MOH (Ministry of Health)**: The government body responsible for national health policy, planning, and services in a country
10. **NTD (Neglected Tropical Disease)**: A group of infectious diseases that primarily affect poor and marginalized populations in tropical and subtropical areas, such as schistosomiasis.
11. **Post-Treatment Survey**: A survey conducted after a treatment campaign (like MDA) to assess the effectiveness of the treatment and the current infection status.
12. **Praziquantel Administration**: The process of distributing and giving praziquantel to individuals or populations, typically as part of a public health program.
13. **Praziquantel Mass Treatment Reported**: Information or data collected about large-scale administration of praziquantel in a community or region, often used for monitoring and evaluation.
14. **Praziquantel**: **Urogenital Schistosomiasis**: A type of schistosomiasis infection caused by *Schistosoma haematobium*, affecting the urinary and genital organs.
15. **Proteinuria**: The presence of abnormal levels of protein in the urine, indicating possible kidney damage, often seen in schistosomiasis patients.
16. **Relative Index of Potential Contamination (RIPC)**: A measure used to estimate how likely a water source or area is to be contaminated with schistosome parasites, based on human and snail activity.
17. **Schistosoma haematobium**: A species of parasitic worm that causes urogenital schistosomiasis, typically found in freshwater sources in Africa and the Middle East.
18. **Schistosome**: A parasitic flatworm of the genus *Schistosoma* that causes schistosomiasis in humans.
19. **Sub-Saharan Africa**: A geographic region of the African continent located south of the Sahara Desert, often discussed in health research due to high disease burden.

## APPENDIX II

### LIST OF ABBREVIATIONS

MDA: Mass Drug Administration
CBS: Community-Based Survey
SBS: School-Based Survey
CDDs: Community Drug Distributors
Rel. IPC: Relative Index of Potential Contamination
NTDs: Neglected Tropical Diseases
WHA: World Health Assembly
IPC: Index of Potential Contamination
MOH: Ministry of Health and Human Services
CDC: Centre for Disease Control and Prevention
WHO: World Health Organization
PTS: Post-Treatment Survey
FCT: Federal Capital Territory

