## Supplementary figures and images for "Index of potential contamination of urogenital schistosomiasis based on mass drug administration of praziquantel among school children in Benue State, Nigeria"

### Ethical Clearance

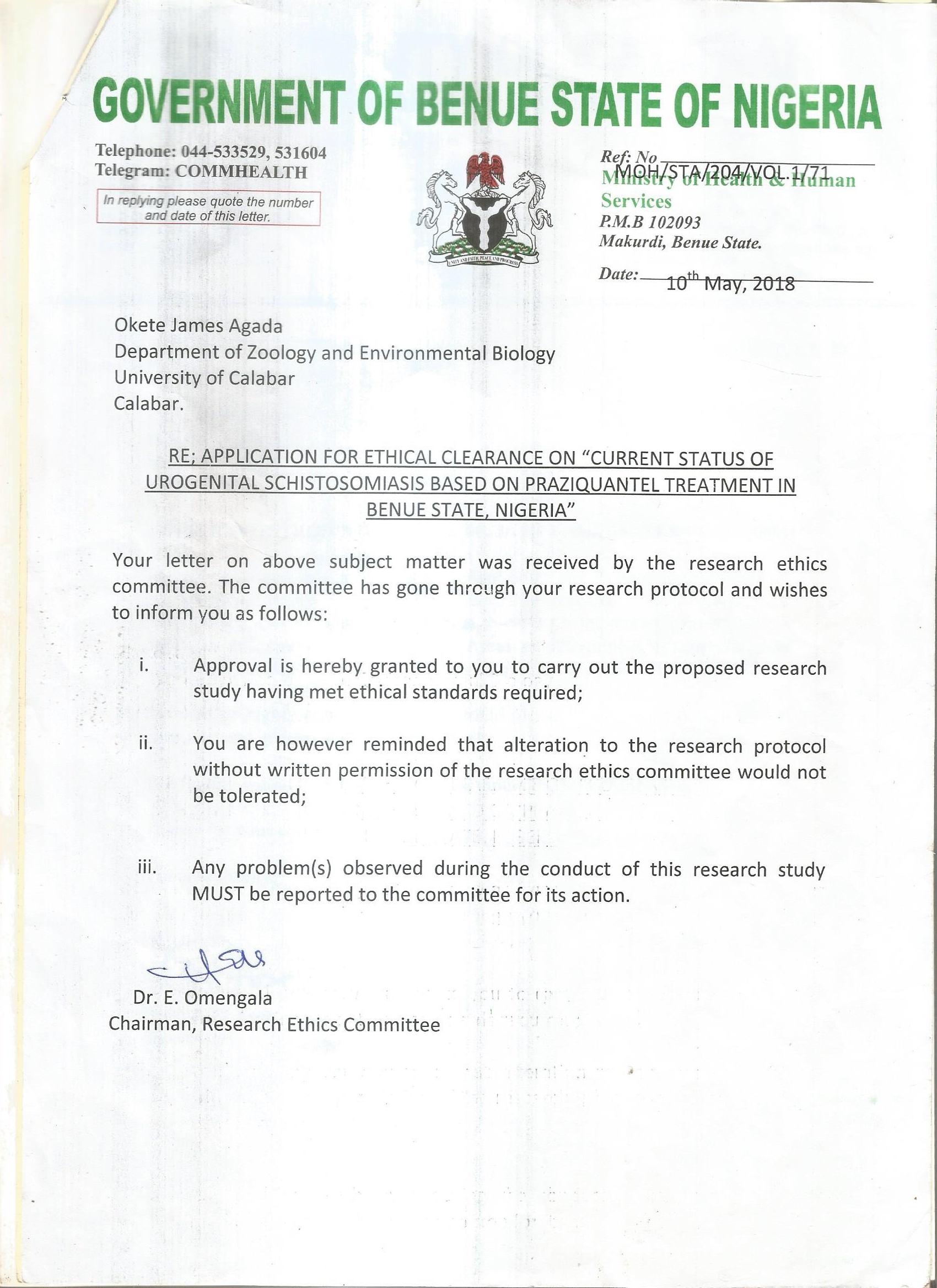
